# Prognostic determinants of anterior large vessel occlusion in acute stroke in elderly patients

**DOI:** 10.1101/2023.08.18.23294293

**Authors:** Takashi Mitsuhashi, Kohsuke Teranishi, Joji Tokugawa, Takumi Mitsuhashi, Makoto Hishii, Hidenori Oishi

**Author notes:** Corresponding Author: Takashi Mitsuhashi, Department of Nerosurgery, Juntendo University Nerima Hospital, 3-1-10 Takanodai, Nerima-ku,Tokyo, Japan.

## Abstract

**Introduction:** Mechanical thrombectomy(MT) has been shown to be safety and effectiveness for acute anterior circulation large vessel occlusion(LVO)of all ages, and together with intravenous thrombolysis has become an standard treatment for acute stroke. In an aging world, the effectiveness of MT for the elderly has not been fully demonstrated. We investigated factors associated with prognosis in elderly patients undergoing MT in Japanese practice, where the elderly are defined as those aged 80 years or older, in the context of an ageing population.

**Method:** MT was performed in 59 cases of LVO of the anterior circulation. Primary outcome evaluated functional outcome at three months. Parametric and/or non-parametric test and a binomial logistic regression model was performed to identify prognosis factors in elderly patients.

**Results:** Overall, Of the 59 patients, 47.5% (28/59) achieved an mRS ≤3 at 3 months. Mortality rate was 20.3% (12/59). Younger age(*P*=0.032), lower NIHSS on admission(*P*=0.00005), lower level of NT-proBNP on admission(*P*=0.041), lower level of D-dimer on admission(*P*=0.01), First Pass Effect (FPE) (*P*=0.024) and good recanalization(*P*=0.0025) were associated with favorable clinical outcome. In the binomial logistic regression model, only lower NIHSS on admission was significantly associated with good clinical outcome.

**Conclusions:** In the present study, not only were younger age, lower NIHSS on admission and FPE already reported as prognostic factors for MT for LVO in the elderly, but also, although not previously reported, lower levels of NT-proBNP on admission and lower level of D-dimer on admission were considered as possible prognostic factors.

## Introduction

Several randomised controlled trials of mechanical thrombectomy (MT) for the treatment of acute anterior circulation large vessel occlusion (LVO) have demonstrated efficacy and safety in all age groups. [1]

MT has become one of the standard treatments for acute ischemic stroke caused by LVO, along with intravenous thrombolysis. Some guidelines recommended considering MT for occlusion of the M1 segment of the middle cerebral artery or internal carotid artery. [2,3] As Hendrix et al. also pointed out, although the guideline proved to be a class 1 recommendation for MT in patients over 80 years of age, several RCTs have excluded patients over 80 or 85 years of age.[4]

On the other hand, it is recognized that life expectancy has increased worldwide in recent years and that the risk of developing ischemic stroke is thought to increase as the number of older people increases and as they get older. Some previous studies have shown that people over 80 years of age have the highest incidence of acute ischemic stroke. [5]

However, the evidence for MT in elderly patients with LVO is scarce and not explicitly stated in the guidelines. Furthermore, Alawieh et al. point out that many of the previous conclusions regarding the efficacy and safety of MT in the elderly were based on single-arm, single-institute studies, resulting in significant heterogeneity among studies and a lack of comparison with optimal medical management. [6]

Previous studies on MT for LVO have highlighted advanced age as a strong predictor of unfavorable outcome after MT. [7,8]

Japan has the highest ageing rate in the world, and other developed countries are also expected to become ageing societies.

The safety and efficacy of MT for LVO have been reported in octogenarians, nonagenarians, and elderly patients with LVO, including prognosis-related factors.

Factors contributing to outcome include successful reperfusion, first pass effect (FPE), low NIHSS score on admission, initial ASPECTS, time from onset to groin puncture, time from groin puncture to recanalization, pre-existing functional deficits, and age over 80 years. [6,9,10,11,12,13]

Although, the HERMES meta-analysis showed that MT had beneficial effects in patients aged 80 years or older patients, the proportional of cases over 80 years of age was low. [1] Therefore, the current guidelines for the United States provide class 1 recommendation for MT in patients aged 80 years or older. [14]

Therefore, Zaidat et al. dichotomized patient age based on previous studies, using 80 years as the cut-off point and defining elderly patients as those aged 80 years or older. In the TRACK registry, age ≥80 years was reported to be a prognostic factor and that the rate of patients with an mRS score 0 to 2 at 90 days decreased slightly with age.[15]

In addition, Meyer et al. pointed out that no substantial therapy recommendation for patients aged 90+ was derived from the recent landmark thrombectomy trials. [16]

In the present study, we defined “elderly” as those over 80 years of age because, as mentioned above, many studies, including previous guidelines, often set the age at which a person is considered elderly at 80 years. Furthermore, since the average life expectancy of Japanese men is in the early 80s and that of women in the mid 80s, we defined those aged 80 and over as elderly.

Since many elderly patients have undergone MT for LVO in our medium-sized hospital in Japan, which has an aging population, we investigated the results of MT for LVO in elderly patients and reviewed the prognostic factors. To identify factors associated with favorable outcome in patients who have underwent MT for LVO.

## Materials and Methods

### Patient selection

Consecutive 141 patients who underwent MT for anterior circulation LVO at our hospital between October 2015 and October 2021 were retrospectively reviewed. In this retrospective single-center study, the protocol was approved by the ethics committee of Juntendo University Nerima Hospital.

The inclusion criteria of the present study were as follows; (1) acute ischemic stroke due to LVO within the anterior circulation, including the internal carotid artery (ICA) or middle cerebral artery (MCA), with or without additional intravenous thrombolysis; (2) age at onset is 80 years or older; (3) known admission National Institute of Health Stroke Scale (NIHSS),Alberta Stroke Program Early CT Score (ASPECTS), and Diffusion-weighted imaging ASPECTS (DWI-ASPECTS) for patients who are eligible for magnetic resonance imaging; (4) known values of the premorbid modified Rankin Scale (mRS) and mRS at 90 days after stroke.

Data were collected on age at admission, sex, time of onset or last known well time, time of arrival at our institute, NIHSS at admission, blood collection data at admission (N-Terminal Pro-Brain Natriuretic Peptide (NT-proBNP) (pg/ml), D-dimer (*μ*g/ml), Glycated hemoglobin (HbA1c) (%)), time of recanalization, number of attempts, and angiographic recanalization status as modified thrombolysis in cerebral infarction (mTICI).

In this study we defined a clinical outcome mRS 0-3 as favorable clinical outcome, for the following reasons.

The mRS is a non-linear scale, a measure of Activity of Daily Life that focuses primarily on mobility, and dichotomization of outcomes has been used in research studies. Many previous studies, including HERMES, have treated mRS0-2 as a good clinical outcome. [1] However, Wilson et al. pointed out that the lower inter-rater reliability in classifying patients as mRS2 and 3 or by the existence of subgroups within the mRS3 group. [17] In Addition, Rangraju et al. described that the patients who achieved mRS3 were more similar to mRS2 than mRS4 patients in terms of on functional disability and quality of life. [18]

The majority of patients underwent MT under local anesthesia with the use of a stent-retriever and/or an aspiration catheter.

### Study End Points

The primary neurological end point of this study was the rate of favourable clinical outcome defined as mRS ≤3 at 90-day follow up, adjusted for patient age.

### Statistical analysis

Variables were compared between two groups: mRS 0-3; favourable clinical outcome and 4-6; unfavourable clinical outcome, using either the t-test or the Mann-Whitney-U test (M-W test), as appropriate, to compared variables between groups. We also developed and analysed a binomial logistic regression model to predict favourable or unfavourable clinical outcomes.

P-values of <0.05 were considered statistically significant. IBM SPSS version 25 (Chicago, IL, USA) was used for data analysis.

## Results

### Patient characteristics

Fifty-nine elderly patients aged over 80 years met the inclusion criteria and were treated between October 2015 and October 2021.

The median age was 85 years (IQR 83-87) and 61.02% (27/59) were female. The median pre-morbid mRS was 0 (IQR0-1), but the mean value was 0.559.

Patients were admitted to hospital with a median NIHSS score of 24 (IQR 18-27), a median baseline ASPECTS score of 9 (IQR 7-9), and a median baseline DWI-ASPECTS score of 7 (IQR 6-8). Six patients were unable to undergo MRI due to metallic implants in their body.

The most common cardiovascular risk factors (Table 1.) were hypertension (83.1%, 49/59), diabetes mellitus (20%, 12/59), hyper lipidemia (13.6%, 8/59) and arterial fibrillation (56%,33/59). There is no statistically significant difference in cardiovascular risk factors between the favourable and unfavourable clinical outcome groups by t-test and M-W test (*p*=0.7802 and *p*=0.7728). The most common site of occlusion (Table 2.) was the middle cerebral artery (MCA), particularly the M1 segment (28.9%,17/59): other site in the anterior circulation included the internal carotid artery (ICA) (22%, 13/59), the MCA M2 segment (18.6%,11/59), the cervical segment of ICA (6.8%, 4/59), and the intracranial segment of ICA (5%, 3/59). There is no statistically significant difference in the site of occlusion between the favourable and unfavourable clinical outcome groups by t-test and M-W test (*p*=0.8129 and *p*=0.7481).

**Table 1.** Patients’ characteristics.

|  | Favourable clinical outcome<br>28 cases | Unfavourable clinical outcome<br>31 cases |
| --- | --- | --- |
| Hypertension | 14 (50%) | 25 (80.65%) |
| Diabetes | 4 (14.29%) | 8 (25.81%) |
| Hyperlipidemia | 5 (17.86%) | 3 (9.68%) |
| Atrial fibrillation | 24 (85.71%) | 19 (61.29%) |

t-test and M-W test ( $p=0.7802$ and $p=0.7728$ )

**Table 2.** Occlusion site. ICA: internal carotid artery, M1:middle cerebral artery horizontal segment, M2/M3: middle cerebral artery insular segment/ opercular segment, tandem: sub-occlusion or stenosis of extracranial internal carotid artery, together with simultaneous intracranial large vessel occlusion.

|  | Favourable clinical outcome<br>28 cases | Unfavourable clinical outcome<br>31 cases |
| --- | --- | --- |
| Cervical ICA | 0 | 4 |
| Intracranial ICA | 1 | 2 |
| Teriminal ICA | 6 | 7 |
| M1 proximal | 6 | 3 |
| M1 distal | 8 | 10 |
| M2/M3 | 7 | 4 |
| Tandem | 0 | 1 |

t-test and M-W test ( $p=0.8129$ and $p=0.7481$ )

More than half patients (61.02%,36/59) underwent intravenous thrombolysis prior to MT.

Base-line characteristics at admission were compared between favourable clinical outcome (mRS ≤3) and unfavourable clinical outcome (mRS >3) at 90days after admission. (table 3.)

**Table 3.** Prognostic factor analysis. CT ASPECTS: Alberta Stroke Early CT score, MRI DWI-ASPECTS: diffusion weighted imaging ASPECTS, NIHSS: National Institutes of Health Stroke Scale, HbA1c: Glycated hemoglobin, NT-proBNP: N-Terminal Pro-Brain Natriuretic Peptide, mTICI: modified Thrombolysis in Cerebral Infarction, t-test: Student’s t-test, M-W test: Mann-Whitney U test, *: statistically significant.

|  | Favorable clinical<br>outcome 28cases | Unfavorable clinical<br>outcome 31 cases | t-stats | *p* | M-W test | *p* |
| --- | --- | --- | --- | --- | --- | --- |
| age, median (IQR) years | 84 (82.75-86) | 85 (83-90) | -2.1998 | 0.03189 * | 329 | 0.1107 |
| female | 11(39.29%) | 16(51.6%) |  |  |  |  |
| CT ASPECTS, median (IQR) | 9 (7-9.25) | 9 (7-9) | -3.6873 | 0.0962 | 502.5 | 0.2867 |
| MRI DWI-ASPECTS, median (IQR) | 7.5 (6-9) | 8 (6-8) | 1.9157 | 0.3511 | 381.5 | 0.587 |
| NIHSS, median (IQR) | 20 (16-24.25) | 26 (22.5-30) | -3.6873 | 0.00051 * | 213 | 0.0008 * |
| intravenous thrombolysis | 18 (64.29%) | 18 (58.06%) |  |  |  |  |
| HbA1c (median, IQR) (%)) | 5.9(2.2-6) | 6(5.625-6.475) | -1.180004 | 0.2431 | 326.5 | 0.211 |
| D-dimer (median, IQR) ($\mu\text{g/ml}$) | 1.25(0.875-1.8) | 2(1.2-4.05) | -0.70065 | 1.4864 | 255 | 0.01037 * |
| NT-proBNP (median, IQR) (pg/ml) | 636.65(314.2-913) | 1195.5(746.6-1814) | -1.6016 | 0.1164 | 169 | 0.04028 * |
| TICI2b < | 26 (92.86%) | 18 (58.06%) | 3.28524 | 0.00175 * | 585 | 0.002457 * |
| number of passes, median (IQR) | 1 (1-2) | 2 (1-2) | 2.34518 | 0.02252 * | 562 | 0.024427 * |
| groin to recanalization time, median (IQR)<br>(min) | 41(21.5-57,25) | 53(36-72) | -1.28237 | 0.20491 | 307 | 0.05475 |
| arrival to recanalization time, median (IQR)<br>(min) | 145(110.75-178.25) | 148(117-177) | 0.816 | 0.4178 | -0.251 | 0.8064 |
| onset to arrival time, median (IQR) (min) | 52(38.75-165.75) | 57(39.5-157) | 1.3385 | 0.186023 | 441 | 0.9214 |

In terms of age, the median age in the favourable clinical outcome group was 84 years (IQR 82.75-86), while the median age in the unfavourable clinical outcome group was 86 years (IQR 83-90), a statistically significant difference found by t-test (*p*=0.0319) (Table.3). In terms of time from onset to arrival, the median time in the favourable clinical outcome group was 52 minutes (IQR 38.75-165.75) and the median time in the unfavourable clinical outcome group was 57 minutes (IQR 39.5-157), with no statistically significant difference by t-test and M-W test (*p*=0.186 and *p*=0.9214) (Table.3). The median NIHSS score on admission was 20 (IQR 16-24.25) in the favourable clinical outcome group and 26 (IQR 22.5-30) in the unfavourable clinical outcome group, a statistically significant difference was found by both t-test and M-W test (*p*=0.0005 and *p*=0.0008) (Table.3). For ASPECTS and DWI-ASPECTS on admission, the median ASPECTS in the favourable clinical outcome group was 9 (IQR 7-9.25) and the median DWI-ASPECTS was 7.5 (IQR 6-9), whereas in the unfavourable clinical outcome group the median ASPECTS was 9 (IQR 7-9) and the median DWI-ASPECTS was 8 (IQR 6-8), and there was no statistically significant difference in either group by t-test and M-W test (*p*=0.0962 and *p*=0.2867, and *p*=0.3511 and *p*=0.587) (Table.3). For the NT-proBNP level (pg/ml) on admission, the median for the favourable clinical outcome group was 636.65 (pg/ml) (IQR 314.2-913), while the median for the unfavourable clinical outcome group was 1195.5 (pg/ml) (IQR 746.6-1814), a statistically significant difference was found by M-W test (*p*=0.0403) (Table.3). For admission D-dimer level (*μ*g/ml), the median for the favourable clinical outcome group was 1.25 (*μ*g/ml) (IQR 1.2-4.05), while the median for the unfavourable clinical outcome group was 2 (*μ*g/ml) (IQR 0.875-1.8), a statistically significant difference was found by M-W test (*p*=0.0104) (Table.3). On the other hand, the median HbA1c level (%) on admission was 5.9 (%) (IQR 5.5-6) in the favourable clinical outcome group and 6 (%) (IQR 5.625-6.475) in the unfavourable clinical outcome group, which was not statistically significant by t-test and M-W test (*p*=0.2431 and *p*=0.211) (Table.3).

In binomial logistic regression analysis with further investigation, only a lower NIHSS score on admission was significantly correlated with a good clinical outcome (*p*=0.010).

Although no significant correlations were found, there was a trend for favourable clinical outcome to correlate with younger age; NT-proBNP showed a slight tendency to correlate with favourable outcome (Table.4).

### Procedural and functional outcomes

These favorable clinical outcomes (mRS ≤3) were observed in 47.5% (28/59) of patients at 90-days after admission. Post-interventional in-hospital mortality was 20.3% (12/59).

Successful angiographic recanalization (mTICI ≥2b) was achieved in 74.6% (44/59) of patients. In contrast, unsuccessful recanalization (mTICI 0) occurred in four cases (6.8%).

The percentage of good recanalization with mTICI 2b or better was 92.9% (26/28) in the favourable clinical outcome group and 58.1% (18/31) in the unfavourable clinical outcome group, a statistically significant differences was found in both t-test and M-W test (*p*=0.0018 and *p*=0.0025) (Table.3).

For the number of attempts, the median for the favourable clinical outcome group was 1(IQR 1-2) and the median for the unfavourable clinical outcome group was 2(IQR 1-2), a statistically significant differences was found in both t-test and M-W test (*p*=0.0225 and *p*=0.0244) (Table.3). For arrival to recanalization time (min), the median for the favourable clinical outcome group was 145 (min) (IQR 110.75-178.25), and the median for the unfavourable clinical outcome group was 140 (min) (IQR 117-177), which was not statistically significant by t-test and M-W test (*p*=0.4178 and *p*=0.8064) (Table.3). For puncture to recanalization time (min), or procedure time, the median for the favourable clinical outcome group was 41 (min) (IQR 21.5-57.25), and the median for the unfavourable clinical outcome group was 53 (min) (IQR 36-72), which was not a statistically significant difference by t-test and M-W test (*p*=0.2049 and *p*=0.054) (Table.3).

Binomial logistic regression analysis showed no significant correlations, but there was a trend for cases with TICI ≥2b and fewer attempts to correlate with favourable clinical outcomes (Table.4).

**Table 4.**
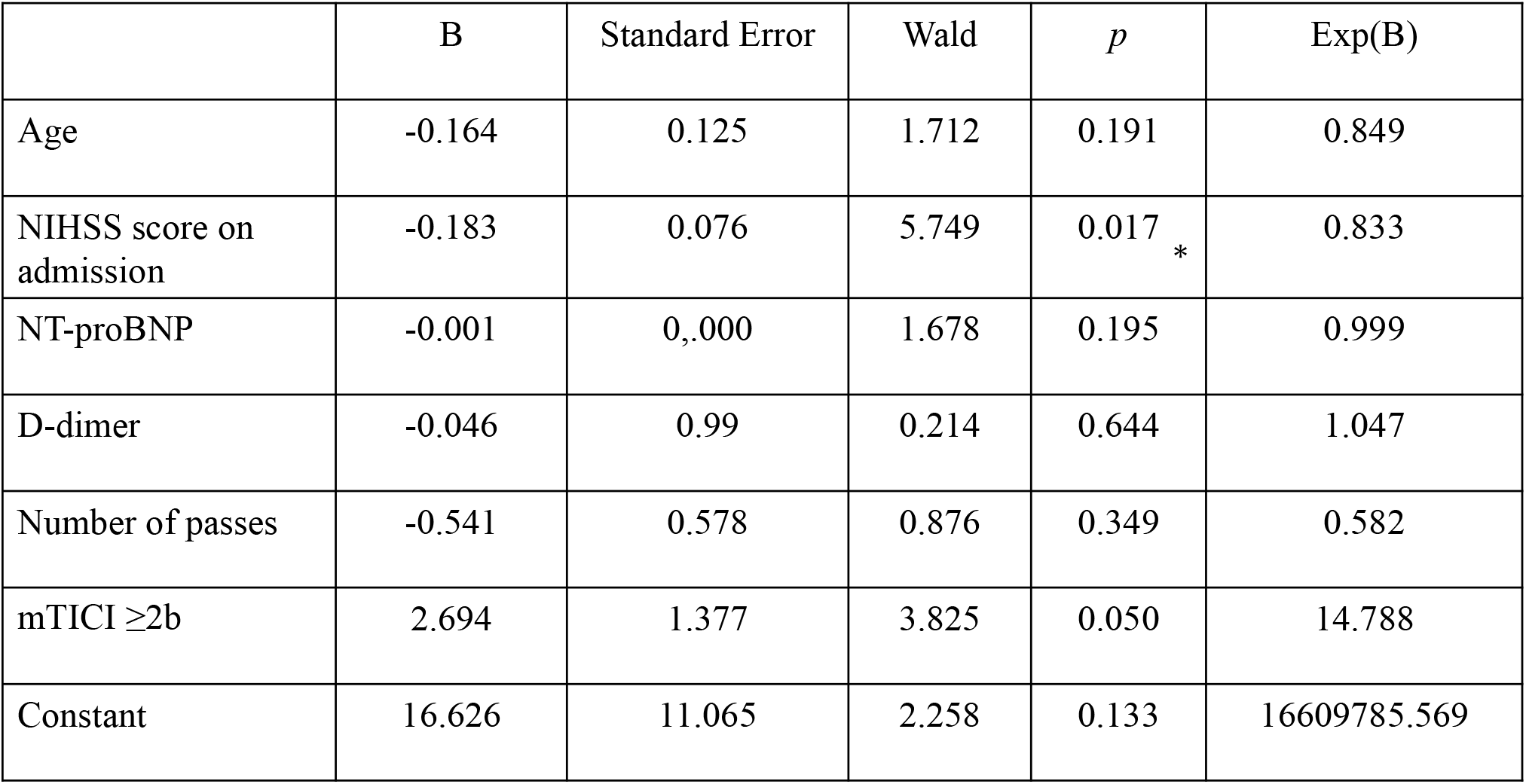
Binomial logistic regression model analysis for prognostic factor. NIHSS: National Institutes of Health Stroke Scale, NT-proBNP: N-Terminal Pro-Brain Natriuretic Peptide, mTICI: modified Thrombolysis in Cerebral Infarction, *: statistically significant.

## Discussion/Conclusion

In this study, in patients over 80 years of age undergoing MT for LVO, the factors associated with a favourable clinical outcome on t-test and M-W test were younger age, lower NIHSS score on admission, lower level of NT-proBNP on admission, lower level of D-dimer on admission, FPE (first pass effect) and good recanalisation, However, binomial logistic regression analysis showed that only a lower NIHSS score on admission was significantly associated with a favourable clinical outcome. For the other factors, only a trend was associated with a favourable clinical outcome.

Based on the hypothesis that many of the younger patients included in many reports of MT outcomes in LVO, including HERMES, had a pre-stroke mRS of 0 [1],

If moving from an mRS of 0 to 2 is considered a favourable clinical outcome, the mean pre-stroke mRS of our study group, consisting of patients over 80 years of age, was 0.559, which is consistent with moving from an mRS of 0 to 3, and we defined an mRS of 0-3 as a favourable clinical outcome.

### Age

In this study, although there were statistically significant differences in t-test, binomial logistic regression analysis showed no significant differences, although it is clear that the older the patient, the unfavourable clinical outcome. Similar findings have been reported previously. [19,20,21] As previously noted, pre-existing physical and/or cognitive disability and the high incidence of complications during hospitalization were thought to limit the potential for post-stroke rehabilitation with increasing age. [16]

In this study of elderly patients with mRS6, deaths from infectious complications such as aspiration pneumonia were more common, and deaths from malignant oedema or intraparenchymal haemorrhagic infarction due to stroke were very rare. In the background, many elderly patients have complications such as frailty, and it is thought that their sputum expectoration and other functions are also declining, so it is necessary to pay attention to infectious complications, especially aspiration pneumonia.

In older patients, atherosclerotic changes in the access route are often significant, and treatment may be discontinued due to access difficulties. If such good recanalization is not achieved, a good clinical outcome is unlikely. [12]

### NIHSS score on admission

In this study, the only factors that were found to be significantly associated between lower NIHSS scores on admission and favourable clinical outcomes were found not only in the t-test and M-W test, but also in binomial logistic regression analysis. Similar findings have been reported previously. [12,22]

Higher NIHSS scores reflect a more extensive cerebral ischaemic state and may indicate ischaemic symptoms due to more proximal occluded arteries; Arsava et al. have noted the presence of poor collateral circulation [23].

### D-dimer

In this study, there was a statistically significant difference between a high level of D-dimer and an unfavorable clinical outcome in M-W test, but not in binomial logistic regression analysis. D-dimer is a degradation product of cross-linked fibrin and a biomarker of the fibrinolytic and coagulation system. [24] Elevated D-dimer levels in acute ischemic stroke have been associated with functional prognosis, acute morbidity, and infarct size. [25,26,27,28]

Hisamitsu et al. described that in patients with acute stroke, elevated D-dimer levels may be related to increased activation of the fibrinolytic system in the thrombus of the occluded artery, and the larger the thrombus, the greater the increase. [29] Furthermore, they speculated that a high D-dimer level leads to an unfavourable clinical outcome because the amount of thrombus in the occluded artery is high, making it difficult to achieve a first pass effect (FPE), resulting in distal embolization and prolonged procedure time.

### NT-ProBNP

BNP is mainly synthesized cardiomyocytes in response to increased cardiac chamber wall stress. [30] Activation of BNP produces an inactive NT-proBNP(N-terminal pro-brain natriuretic peptide). [31] NT-proBNP has a longer half-life than BNP and results in less fluctuation, but has been reported to be associated with adverse clinical outcomes. [32,33,34] Plasma NT-proBNP levels have been reported to be elevated in patients with heart failure, left ventricular dysfunction, acute coronary syndromes, and cardiogenic stroke.[35,36] In addition, NT-pro BNP is a biological marker of cerebrovascular disease for identifying ischemic stroke subtypes, and predicting the incidence of arterial fibrillation related stroke. [37,38] In this study, almost all patients with elevated plasma NT-proBNP had a cardiogenic embolism. Furthermore, there is increasing evidence that serum levels of NT-proBNP are associated with an increasing risk of adverse clinical outcome in patients after stroke. [33,39]

There are no reports on the association between NT-proBNP and prognosis in MT for LVO in the elderly patients. Chronic heart failure has been reported to exacerbate cerebrovascular disease, dementia, and pneumonia and to reduce cardiac activity. [40,41,42]

In this study, although there were statistically significant differences in the association of FPE with favourable clinical outcomes in both t-test and M-W tests, there were no significant differences in binomial logistic regression analysis. The results suggesting that cardiac function also contributes to improved prognosis after rehabilitation. Therefore, cardiac function may be an indicator of functional prognosis after MT for LVO.

In this study, NT-proBNP levels were elevated well above 150 ng/ml, which is considered a marker for heart failure. However, most patients had no obvious symptoms of heart failure. In a case-cohort study from the REGARDS cohort, the authors confirmed that the association of NT-proBNP with stroke was stronger for cardioembolic stroke. [43]

Zhang et al. proposed that three possible mechanisms of NT-pro BNP elevation; ➀ central blood vessels damage promotes massive release of NT-proBNP from the CNS to the blood, resulting in an absolute increase of NT-proBNP, ➁ NT-proBNP concentrations derived from the same precursor protein increase simultaneously when cerebral infarction occurs; however, because of their different patterns of metabolism, the activation of the endocrine system controlled by CNS degrade BNP more than NT-proBNP, resulting in a relative increase of NT-proBNP, ➂ following the deterioration that occurs due to heart failure, organisms begin to mobilize activity factors that collaborate with the rennin-angiotensin-aldosterone system to induce a diastolic state in blood vessels. [44]

In this study, the mechanism of NT-proBNP level elevation in LVO is not only one of the above three possibilities, but also the result of a sudden increase in blood pressure to promote spontaneous recanalization of the main artery occlusion, which may have resulted in an excessive cardiac load.

The cases with elevated level of NT-proBNP and unfavorable clinical outcome included those who died due to complications such as heart failure or aspiration pneumonia during the course of hospitalization. Some cases were already complicated by aspiration pneumonia at the onset of hospitalization, which may have played a role in the elevated NT-proBNP levels.

Many of the cases with elevated level of NTproBNP are consequently older with higher NIHSS, suggesting that more severe and more extensive fast running has occurred since then, and the elevation may be the result of greater sympathetic activation

In the present study, the regression analysis failed to produce significant results due to insufficient power of nunber. Therefore, if n is increased, the regression model may be able to produce results showing that NPproBNP is an independent predictor of prognosis.

### FPE

In this study, although there were statistically significant differences in both t-test and M-W test, binomial logistic regression analysis showed no significant differences. Zaidat et al. showed that patients with mTICI 3 had better clinical outcome, lower mortality and fewer procedural adverse events after one pass.[45] Rousiers ED et al. described that FPE is a significant determinant of good clinical outcome in the oldest patients. Similar effects were observed in our results. [18] FPE has been shown to reduce procedure time, ischemic core volume expansion, and the incidence of procedural complications. [46,47] However, our results on puncture to recanalization time showed no correlation with clinical outcome.

## Limitations

Nevertheless, the present study has some limitations. The study design is retrospective and non-randomized with a small sample size treated at a single institution. We did not compare patients treated with MT with those treated medical treatment including thrombolysis alone. We did not perform serial measurement of NT-proBNP levels and D-dimer levels; therefore, we were unable to examine the association of changes in these levels during after acute stroke phase. In the future, patient data should be accumulated and a large-scale prospective study should be conducted. Although the regression analysis could not produce significant results due to the insufficient power of the number, a certain trend can be observed, so if the number is increased in the future, it may be possible to produce results of an independent prognostic predictor in a regression model.

## Conclusion

The study found that factors associated with favourable clinical outcomes in patients aged 80 years or older undergoing MT for LVO were younger age, lower NIHSS score on admission, lower level of NT-proBNP on admission, lower level of D-dimer on admission, FPE, and better recanalisation, with a strong correlated with lower NIHSS on admission, but a trend was observed for other factors. There are no reports on the correlation between NT-proBNP levels on admission and favourable clinical outcomes in MT for LVO in the elderly, and further verification is needed.

## Acknowledgments

The medical assistants at Juntendo Nerima Hospital, Miyuki Yanagi and YokoTanaka, are thanked for their cooperation in data collection..

## Statement of Ethics

This retrospective single-center study, the protocol was approved by the Ethics Review Board at Juntendo University Nerima Hospital (18-39).

## Author Contributions

Takashi Mitsuhashihad full access to all of the data in the study and take responsibility for the integrity of the data and the accuracy of the data analysis. Concept and design: Takashi Mitsuhashi, Kohsuke Teranishi, Joji Tokugawa, Makoto Hishii, and Hidenori Oishi. Acquisition of data: Takashi Mitsuhashi. Drafting of the manuscript: Takashi Mitsuhashi, Joji Tokugawa, and Takumi Mitsuhashi. Critical revision of the manuscript for important intellectual content: Kohsuke Teranishi and Hidenori Oishi. Obtained funding: Takashi Mitsuhashi and Joji Tokugawa.

## Data Availability Statement

The data that support the findings of this study are not publicity available due to their containing information that could compromise the privacy of research participants but are available from the corresponding author upon reasonable request.

